# Delineation and monitoring of the T cell repertoire of adoptive cell transfer product during the treatment of advanced melanoma

**DOI:** 10.1101/2025.01.17.24311574

**Authors:** Cameron Kerr, Shirin Soleimani, David T. Mulder, Arash Nabbi, Diana Gray, Stephanie Pedersen, Valentin Sotov, Sumedha Sudhaman, Linh Nguyen, Naoto Hirano, Pamela S. Ohashi, Marcus O. Butler, Trevor J. Pugh

**Affiliations:** Princess Margaret Cancer Centre, University Health Network, Toronto, Ontario, Canada; University of Toronto, Toronto, Ontario, Canada; Ontario Institute for Cancer Research, Toronto, Ontario, Canada

**Keywords:** T cell repertoire sequencing, tumour infiltrating lymphocytes, adoptive-cell transfer

## Abstract

**Background:** Adoptive cell transfer (ACT) of Tumour-infiltrating lymphocytes (TIL) is an investigational treatment for solid tumours, with preliminary results showing objective clinical responses in some metastatic melanoma patients. The ability to sequence and track the T cell repertoire throughout ACT of TILs provides a method to identify T cell repertoire features associated with patients’ benefit from ACT. Identification of response biomarkers for patients receiving ACT of TILs has been limited. Conflicting evidence is observed in biomarkers such as the number of TILs in the infusion product, with some studies suggesting a relationship with response and others not. Meanwhile, certain potential biomarkers, such as the diversity of the post-infusion peripheral repertoire, have not yet been studied.

**Methods:** In this study, we sought to determine 1) the efficacy of using CapTCR-seq to track TILs in serial blood draws over the course of ACT immunotherapy 2) whether peripheral T cell repertoire statistics are associated with ACT response. In this study, 9 patients with cutaneous (n = 7) or mucosal (n = 2) melanoma received TIL ACT after chemotherapeutic depletion. Hybrid-capture CapTCR-seq was conducted on pre-/post-transfer peripheral blood mononuclear cells (PBMC) and cell-free DNA.

**Results:** Comparison between PBMC DNA, PBMC RNA, and circulating free DNA (cfDNA) repertoires demonstrated an increased presence of shared T cell clonotypes post-infusion when compared with baseline samples. Higher abundance of TIL clonotypes in the PBMC baseline and post-infusion DNA T cell repertoires and the presence of shared DNA T cell clonotypes between timepoints was seen in responders when compared with non-responders according to RECIST criteria.

**Conclusion:** These results demonstrate effective tracking methodologies and suggest a predictive role for baseline repertoire statistics in response to the ACT of TILs.

**Trial registration:** University Health Network Research Ethics Board #11-0683

## 1. BACKGROUND

Adoptive Cell Transfer (ACT) of expanded tumour infiltrating lymphocytes (TILs) has been an effective treatment for multiple types of solid tumours, with complete responses demonstrated in melanoma [3, 6, 8, 17, 19, 23], breast cancer [30], lung cancer [5], cervical cancer [24], and bile duct cancer [26]. Despite cytotoxic capacity of dominantly effector population of T cells in patient-derived product, poor persistence and rapid terminal exhaustion of infused T lymphocytes result in diminished clinical efficacy of ACT [1, 2, 28]. Immune checkpoint blockade therapy as well as immunodepletion followed by TIL ACT present the opportunity to reactivate these exhausted TILs.

ACT immunotherapy starts with preparative host refinement that entails a lymphodepleting regimen (chemotherapy in the current study) that aims 1) to eliminate regulatory T cells and suppressor myeloid populations to prevent potential interference with effector function of transferred anti-tumour T cells, and 2) to limit the population of endogenous cells competing for cytokines. In combination with ACT of expanded TILs and immune checkpoint blockade therapy, the treatment disrupts the endogenous T cell population [22, 27]. Accurate and longitudinal molecular monitoring over the patient timeline during ACT of TILs is a necessary next step for the development of the treatment.

Significant attention has been directed towards studying characteristics of the infusion product and exploring associations with response according to RECIST and irRC criteria. Previous studies involving the ACT of TILs after lymphodepleting chemotherapy have examined the relationship between the number of TILs administered and patient response. While some studies find a positive relationship between the two [3, 8, 12, 19], other have found no relationship [14, 20, 23]. Studies examining the fraction of the infusion product consisting of CD8^+^ T cells in relation to patient response to ACT of TILs after lymphodepleting chemotherapy present similar conflicting results [3, 8, 14, 19, 20, 23]. Studies analysing the relationship between tumour antigen reactivity of the infusion product and patient response during ACT of TILs after lymphodepleting chemotherapy have also provided mixed results, some demonstrating a positive association [5, 24] and some finding no association [19, 20]. It has also been found that the diversity of the TIL infusion is not associated with response in patients given ACT of TILs after lymphodepleting chemotherapy [14].

Studies analysing the post-infusion T cell repertoire have done so through the isolation of T cells from peripheral blood mononuclear cells (PBMC). A long-standing statistic for analysing post-infusion samples is to calculate persistence; the percentage of a post-infusion sample taken up by T cells present in the infusion. Using this metric, increased persistence of T cells from the infusion product present in the post-infusion PBMCs is associated with increased response as determined by RECIST criteria [14, 20, 23]. Despite polyclonal infusions, persistence has been demonstrated to be the result of immunodominant clonotypes [4, 6, 11, 20, 26]. Individual immunodominant Vβ families have been shown to persist for up to 16 months post-infusion with clonotypes that recognize tumour antigens [6, 11]. The presence of immunodominant clonotypes taking up >5% of the peripheral T cell repertoire post-infusion is associated with tumour regression in patients receiving ACT of TILs with prior lymphodepleting chemotherapy [20]. Similarly, complete responses in patients receiving ACT of tumour-specific T cells from peripheral blood were associated with the expansion of single clonotypes with longer half-times in the peripheral repertoire [4].

Our objective in this study was to determine the efficacy of using CapTCR-seq to track TILs in serial blood draws over the course of ACT immunotherapy. Additionally, we sought to determine whether peripheral T cell repertoire statistics are associated with ACT response.

## 2. METHODS

### 2.1 Probe Design

We utilised a modified version of the published CapTCR-seq protocol that utilised three-step J capture/depletion/V capture approach. Briefly, all annotated V (V-panel), D, J (J panel) gene segments and V 3’-UTR (depletion panel) sequences were retrieved from the IMGT / LIGM-DB website (www.imgt.org 9). The 100bp of annotated 3’ V gene coding regions, up to 100 bp, when available, of annotated 5’ J gene coding regions, and 120bp of V 3’-UTR sequences were selected as baits. Probes with duplicate sequences were not included. The V-panel consists of 299 probes (IDT) targeting the 3’ and 5’ 100bp of all TR V gene regions, and the J-panel consists of 95 probes targeting the 5’ 100bp of all TR J gene regions as annotated by IMGT (four loci, 1.8Mb, total targeted 36kb). The depletion-panel consists of 131 probes targeting the 5’ 120bp of 3’-UTR Immunoglobulin V regions, and 107 probes targeting the 5’ 120bp of 3’-UTR TCR V regions.

### 2.2 DNA isolation

CD3+ T cells were isolated by flow assisted cell sorting of PBMC populations separated from whole blood. Peripheral blood mononuclear cells (PBMC) were isolated from whole blood by centrifugation followed by DNA isolation with a Gentra Puregene kit (Qiagen) according to the manufacturer’s protocol. In the case of fresh/frozen tissues, a Qiagen Allprep kit (Qiagen) was employed to extract DNA and RNA, according to the manufacturer’s instructions. The whole blood plasma fraction was then treated with a red blood cell lysis buffer and circulating DNA (cfDNA) was extracted using the Qiagen Nucleic Acid kit (Qiagen) according to the manufacturer’s protocol.

### 2.3 cDNA synthesis

mRNA was separated from isolated total RNA using the NEBNext Poly(A) mRNA Magnetic Isolation Module (NEB) according to manufacturer’s instructions. To generate cDNA, first NEBNext RNA First Strand Synthesis Module (NEB) was used followed by NEBNext RNA Second Strand Synthesis Module (NEB) according to manufacturer’s instructions.

### 2.4 Library preparation

Isolated genomic DNA or synthesised cDNA was diluted in TE buffer to 130uL volumes. Shearing to ∼275 bp was then performed on either a Covaris M220 Focused-ultrasonicator or E220 Focused-ultrasonicator, depending on sample throughput, with the following settings: for a sample volume of 130 μL and desired peak length of 200 bp, Peak Incident Power was set to 175 W; duty factor was set to 10%; cycles per burst was set to 200; treatment time was set to 180 s. In addition, temperature and water levels were carefully held to manufacturer’s recommendations given the instrument in use.

Illumina DNA libraries were generated from 100 – 1000 ng of fragmented DNA using the KAPA HyperPrep Kit (Sigma) library preparation kit following manufacturer’s protocol version 5.16 employing NEXTFlex sequencing library adapters (BIOO Scientific). Library fragment size distribution was determined using the Agilent TapeStation D1000 kit and quantified by fluorometry using the Invitrogen Qubit.

### 2.5 Hybrid capture

For cDNA derived libraries, hybridization was performed with a pooled panel of probes targeting V and J loci in equimolar concentrations. For genomic DNA derived libraries, hybridization and capture was performed iteratively with probes specifically targeting V loci, 3’-UTR sequences, or J loci under standard SeqCap (Roche) conditions with xGen blocking oligos (IDT) and human Cot-1 blocking DNA (Invitrogen). Hybridization is performed at 50 C overnight. The Capture process consisting of bead incubations and washes is performed at 50C.

For the iterative hybridization and capture process, the first J hybridization and capture is performed in completion with terminal PCR amplification with 4 steps. Following clean-up by Agencourt AMPure XP SPRI bead purification (Beckman) this product is used as input for a subsequent depletion step. For depletion, a modified and truncated SeqCap protocol is employed wherein following incubation of the hybridization mixture with M-270 streptavidin linked magnetic beads (Invitrogen), the 15uL hybridization reaction is separated on a magnetic rack, the supernatant is recovered and diluted to 100uL with TE buffer, followed by clean up by standard Agencourt AMPure XP SPRI bead purification (Beckman). The depletion-probe-target-beads are discarded. The purified supernatant is then used as input for a subsequent V-panel capture and hybridization as described above, but with terminal PCR amplification with 16 or amplification steps to achieve sufficient library for sequencing.

### 2.6 Capture analysis

A custom pipeline was developing using Bash/Python/R for the analysis of paired-read sequencing data from Illumina NextSeq 2500 of the hybrid-capture products. The sequencing data was processed using MiXCR-2.1.12 to identify and sort through defined Complementary Determining Region 3 (CDR3) sequences. MiXCR was run using RNA mode for genomic samples to leverage increased alignment capabilities provided by this setting. Partial alignments were allowed, and the resulting TCR and BCR chains were exported to individual clone files.MiXCR log files were employed for quality control of the samples. Specific cutoff numbers were established for each sample cohort. Genomic DNA samples were downsampled to 2,500,000 total sequencing reads, cDNA samples to 170,000 total sequencing reads, and cfDNA samples to 1,000,000 total sequencing reads. An exception was made for the baseline gDNA sample for TLML 4, which had a total of 1,901,038 total sequencing reads. The saturation level for this sample was deemed comparable to other gDNA samples. All downsampling was performed using the subsampling tool of the seqtk package.

### 2.7 Quality control

To assess the depth of sequencing for each sample, we investigated the relationship between the number of TCR clonotypes and sequencing reads. Downsampling was performed to create 18 copies of the original sample, with the total sequencing reads decreasing in 5% increments from the initial downsampling point. The resulting data were used to construct a generalized additive model (GAM) that visualised the relationship between sequencing reads and TCR clonotypes. The first derivative of this model approaching zero indicates that increasing the number of sequencing reads would not yield new TCR clonotypes. Although the first derivative did not reach zero for all models, we observed that the 95% confidence intervals overlapped at the last observation in the model. Consequently, it was determined that all samples were sequenced to a comparable depth. Furthermore, quality control included the assessment of unproductive clones, which revealed that gDNA samples consisted of 22.1% unproductive clones. As a result, these clones were removed from subsequent analysis, along with the exclusion of singletons.

### 2.8 Repertoire analysis

Further repertoire analysis focused on the Beta chain of the TCR. For measuring diversity, multiple indices were used to gain a collectively exhaustive view. Overall diversity was measured using both the Simpson’s and Shannon’s diversity indices. Simpson index: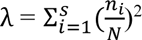, Shannon index: *H* = 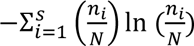.

The inverse Simpson’s index was used to represent diversity and calculated as 1/. For both indices, *n_i_* represents the number of clones in the *i*th clonotype and *N* represents the total clones in all clonotypes. Shannon’s diversity index incorporates both the richness and evenness of the T cell repertoire. Richness of the T cell repertoire was calculated as the total number of clonotypes present in the repertoire is represented as *S*. Shannon evenness index: 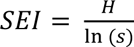, Clonality: *C* = 1 − *SEI*. The Morisita’s overlap index was used to represent the similarity between two T cell repertoires. X and Y were the total number of clones in separated samples, *x_i_* and *y_i_* represented the number of clones present in clonotype *i*. Morisita’s overlap index: 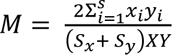.

Risk analysis was used to provide a measure of the risk of an event (presence in the expanding section of a specified sample) among individuals exposed to a particular factor (presence in a specific reference sample) compared to those not exposed. The risk ratio was calculated by dividing the proportion of expanding clones among the clonotypes present in the reference sample by the proportion of expanding clones among the clonotypes not present in the reference sample. Relative risk: 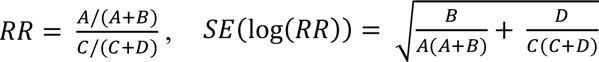

|  | Presence in expanding<br>clonotypes of specified sample | Presence in background clonotypes<br>of specified sample |
| --- | --- | --- |
| Presence in alternate sample | *A* | *B* |
| Absence in alternate sample | *C* | *D* |

A relative risk ratio greater than 1 represents a higher risk for a clonotype to be in the expanding section if it is in the reference sample. To estimate the precision of the risk ratio, we calculated the 95% confidence interval (CI). If the risk ratio’s 95% CI contains 1 it was determined that presence in the reference sample didn’t affect presence in the expanding section. For further information on calculations and visualisations refer to the GitHub page for this study (https://github.com/pughlab/CapTCR-TIL-Tracking).

## 3. RESULTS

### 3.1 Study Design

The protocol enrolled 9 subjects with cutaneous (n = 7) or mucosal (n = 2) melanoma from 2013 to 2016. Before the chemotherapeutic immune depletion, blood and tumour biopsies were collected (Figure 1A). The patients were then administered chemo depletion and a successive expanded TIL-infusion product [15]. Additional serial blood draws were taken every 27 - 106 days after the infusion. The number of post-infusion peripheral blood samples varied from 1 - 17. Patients were grouped according to their response evaluation separated into partial response (PR), stable disease (SD), or progressive disease (PD). For statistical analysis, patients were simplified into responders (PR) and nonresponders (SD and PD) (Figure 1C). Patient IDs used in this paper are de-identified and not known outside the research group.

**Figure 1.**
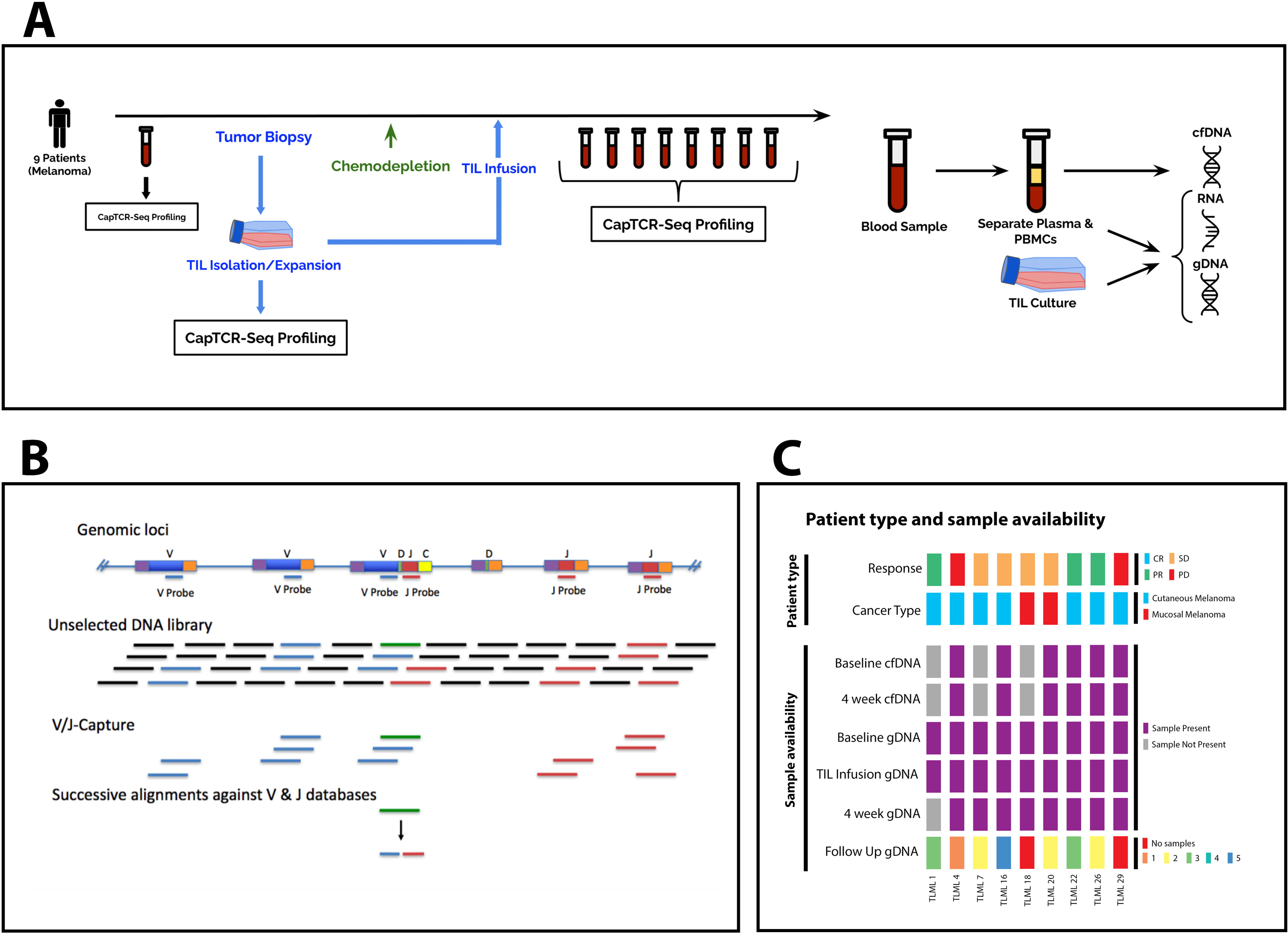
Overview of study design, samples and hybrid capture protocol. (A) 9 melanoma patients underwent the adoptive-cell transfer of TILs with a prior lymphodepleting chemotherapy regimen. Peripheral blood samples were taken pre- and post-infusion and underwent subsequent hybrid-capture profiling using CapTCR-seq. (B) CapTCR-seq schematic; V-regions are coloured blue, J-regions coloured red, D-regions coloured green, constant regions coloured yellow, and non-TCR coding regions coloured black. (C) Patient type and sample availability. Response according to RECIST criteria. Baseline, sample 4 weeks after infusion, and follow-up gDNA samples are from peripheral blood mononuclear cells.

### 3.2 Cross-sample-cohort correlation and overlap increases post-infusion

Genomic DNA, RNA, and cfDNA represent different conceptual aspects of the T cell repertoire. To investigate how differences in sample type impact the ability to track and monitor the T cell repertoire over time we quantified the diversity, richness, and repertoire overlap of repertoires in different sample types. Genomic DNA, RNA, and circulating free DNA (cfDNA) T cell repertoires had different levels of diversity (Figure 2C). Genomic DNA samples had a median inverse Simpson’s diversity of 54.51 (min: 4.41, max: 805.24, n = 44) and a median richness of 513.5 (min: 45, max: 1814, n = 44). RNA samples had a median inverse Simpson’s diversity of 81.78 (min: 5.67, max: 2097.38, n = 39) and a median richness of 2467 (min: 533, max: 7673, n = 39). cfDNA had a median inverse Simpson’s diversity of 16.39 (min: 9.89, max: 38.05, n = 12) and a median richness of 29 (min: 14, max: 163, n = 12). A one-way ANOVA on the log-10 transformed data was performed in order to determine the relationship between sample cohorts and their inverse Simpson diversity. It was found that DNA, RNA, and cfDNA present statistically different levels of diversity (F = 11.964, p = 0.000049). According to Tukey’s HSD multiple comparison test, a significant difference was found between DNA-cfDNA (p = 0.031) and RNA- cfDNA (p = 0.0030) yet no difference was found between DNA-RNA (p = 0.41).

**Figure 2.**
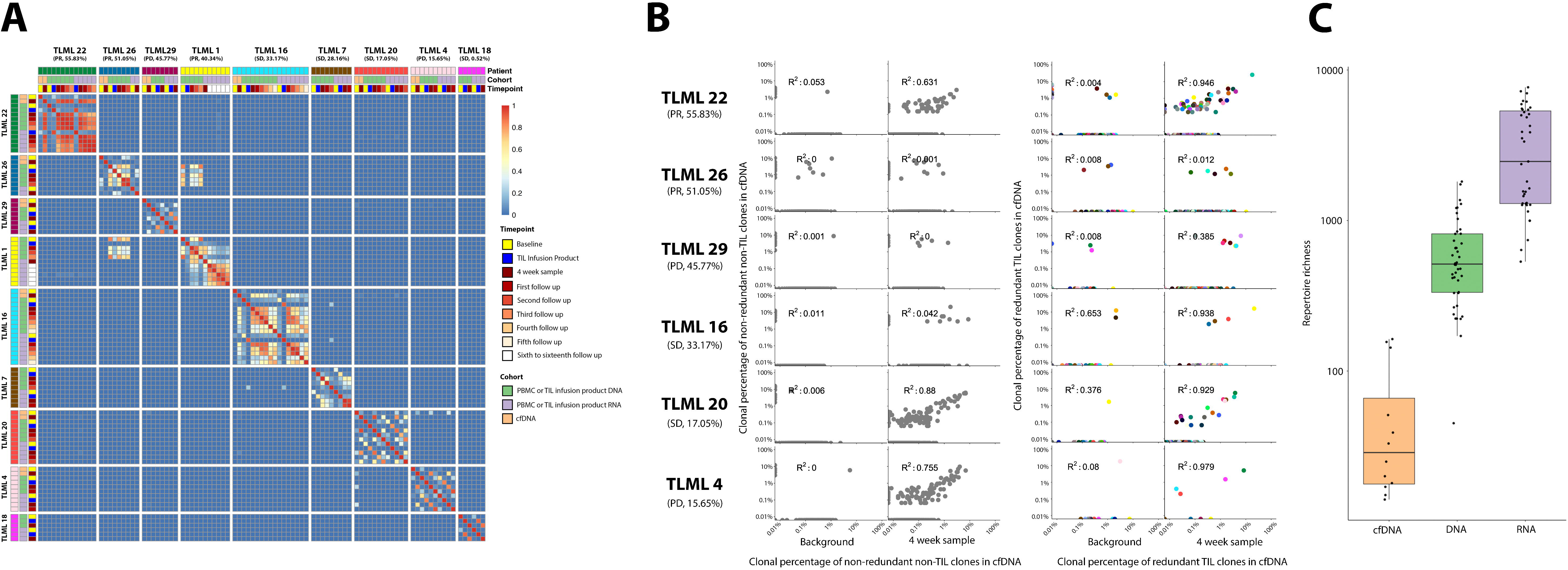
T cell repertoire overlap and correlation across sample cohorts and timepoints. Patients are sorted according to the abundance of tumour-infiltrating lymphocyte clonotypes in the first post infusion peripheral blood mononuclear cell DNA repertoire, as shown below each patient ID along with response according to RECIST criteria. (A) Heatmap of Morisista’s overlap index between all samples. Patients are sorted according to the abundance of tumour-infiltrating lymphocyte (TIL) clonotypes in the first post-infusion peripheral blood mononuclear cells (PBMC) DNA repertoire. (B) Scatter plots displaying correlation between a clonotype’s abundance in the circulating free DNA (cfDNA) and PBMC DNA repertoires. Clonotypes are coloured identically to figure 2B. (C) Boxplot displaying differences in richness of cfDNA, DNA, and RNA. DNA and RNA repertoires include both PBMC and TIL infusion product samples.

To compare the genomic DNA, RNA, and cfDNA T cell repertoires, we calculated the cross-sample- cohort correlation and Morisita’s overlap index for each unique sample (Figure 2A). The cross-sample- cohort correlation measures the similarity of clonotype abundance between DNA, RNA, and cfDNA samples, indicating whether the proportion of the T cell repertoire occupied by each clonotype is consistent across the sample-cohorts.

RNA samples exhibited a median Morisita’s overlap index of 0.84 with their respective DNA sample across all timepoints (min: 0.05, max: 0.95, n = 33). DNA and RNA samples demonstrated a cross-sample-cohort correlation with a median R^2^ value of 0.74 (min: 0.016, max: 0.96, n = 33) across all timepoints. Circulating free DNA demonstrated a median Morisita’s overlap index of 0.13 with their respective DNA sample over all timepoints (min: 0.0041, max: 0.94, n = 12). DNA and cfDNA demonstrated a weak cross-sample-cohort correlation with a median R^2^ value of 0.044 (min: 0.000040, max: 0.92, n = 12) across all timepoints.

To demonstrate the molecular effects of ACT of TILs on different sample cohorts, we calculated the cross-sample-cohort correlation and Morisita’s overlap index for baseline and the first post-infusion sample. The overlap between sample-cohorts was shown to increase post-infusion. PBMC baseline RNA and DNA samples demonstrated a median Morisita’s overlap index of 0.44 (min: 0.20, max: 0.65, n = 6) while the first post-infusion PBMC RNA and DNA samples demonstrated a median Morisita’s overlap index of 0.86 (min: 0.56, max: 0.91, n = 6). The higher post-infusion overlap was determined to be significant through a paired two-tailed t-test (p = 0.0061). Similarly, baseline cfDNA and PBMC DNA samples demonstrated a median Morisita’s overlap index of 0.062 (min: 0.0041, max: 0.18, n = 6) while the first post-infusion cfDNA and PBMC DNA samples demonstrated a median Morisita’s overlap index of 0.70 (min: 0.056, max: 0.94, n = 6). The higher post-infusion overlap was determined to be significant through a paired two-tailed t-test (p = 0.014).

Each PBMC RNA and cfDNA repertoire was subsequently split between clonotypes which are present in the TIL infusion product and at least one PBMC sample and those which are not. The cfDNA- DNA and RNA-DNA cross-sample-cohort correlations were evaluated for clonotypes in each clonotype group (Figure 2B). TIL clonotypes demonstrated no cfDNA baseline cross-sample-cohort correlation with a median R^2^ value of 0.044 (min: 0.0038, max: 0.65, n = 6) and non-TIL clonotypes demonstrated no cfDNA baseline cross-sample-cohort correlation with a median R^2^ value of 0.0033 (min: 0.000054, max: 0.053, n = 6). Similar results were seen in the PBMC RNA baseline samples with a median R^2^ value of 0.65 (min: 0.0070, max: 0.997, n = 6) for TIL clonotypes and 0.10 (min: 0.021, max: 0.28, n = 6) for non-TIL clonotypes.

Post-infusion samples demonstrated an increased cross-sample-cohort correlation as expected, yet only post-infusion TIL clonotypes demonstrated a strong cross-sample-cohort correlation. TIL clonotypes demonstrated a post-infusion cfDNA-DNA cross-sample-cohort correlation with a median R^2^ value of 0.93 (min: 0.012, max: 0.98, n=6) while non-TIL clonotypes demonstrated a post-infusion cross-sample- cohort correlation with a median R^2^ value of 0.34 (min: 0.00020, max: 0.88, n=6). Similar results were seen in the RNA post-infusion samples with a median RNA-DNA cross-sample-cohort correlation with an R^2^ value of 0.77 (min: 0.053, max: 0.97, n = 6) for TIL clonotypes and 0.42 (min: 0.048, max: 0.85, n = 6) for non-TIL clonotypes. The increased correlation of TIL clonotypes between cfDNA-DNA and RNA-DNA was confirmed using paired two-tailed t-tests (cfDNA: p = 0.015, RNA: p = 0.012).

Overall, the overlap between PBMC DNA, PBMC RNA, and cfDNA samples was low at baseline. However, overlap across samples nearly doubles post-infusion. Additionally, a higher correlation was demonstrated between TIL clonotypes when compared with non-TIL clonotypes, demonstrating a more consistent representation of TIL clonotypes across cfDNA, PBMC DNA, and PBMC RNA samples.

### 3.3 Abundance of TILs in baseline and post-infusion PBMC DNA repertoires is associated with response

In previous studies, the persistence of TILs post-infusion has been demonstrated to be associated with clinical response to ACT [14, 20, 23]. To investigate the importance of persistence in relation to clinical response, the abundance of TIL clonotypes in both the baseline and first post-infusion samples was calculated (Figure 3). TIL abundance was defined as the proportion of the T cell repertoire at a certain time point occupied by TIL clonotypes, clonotypes present in the DNA TIL infusion product.

**Figure 3.**
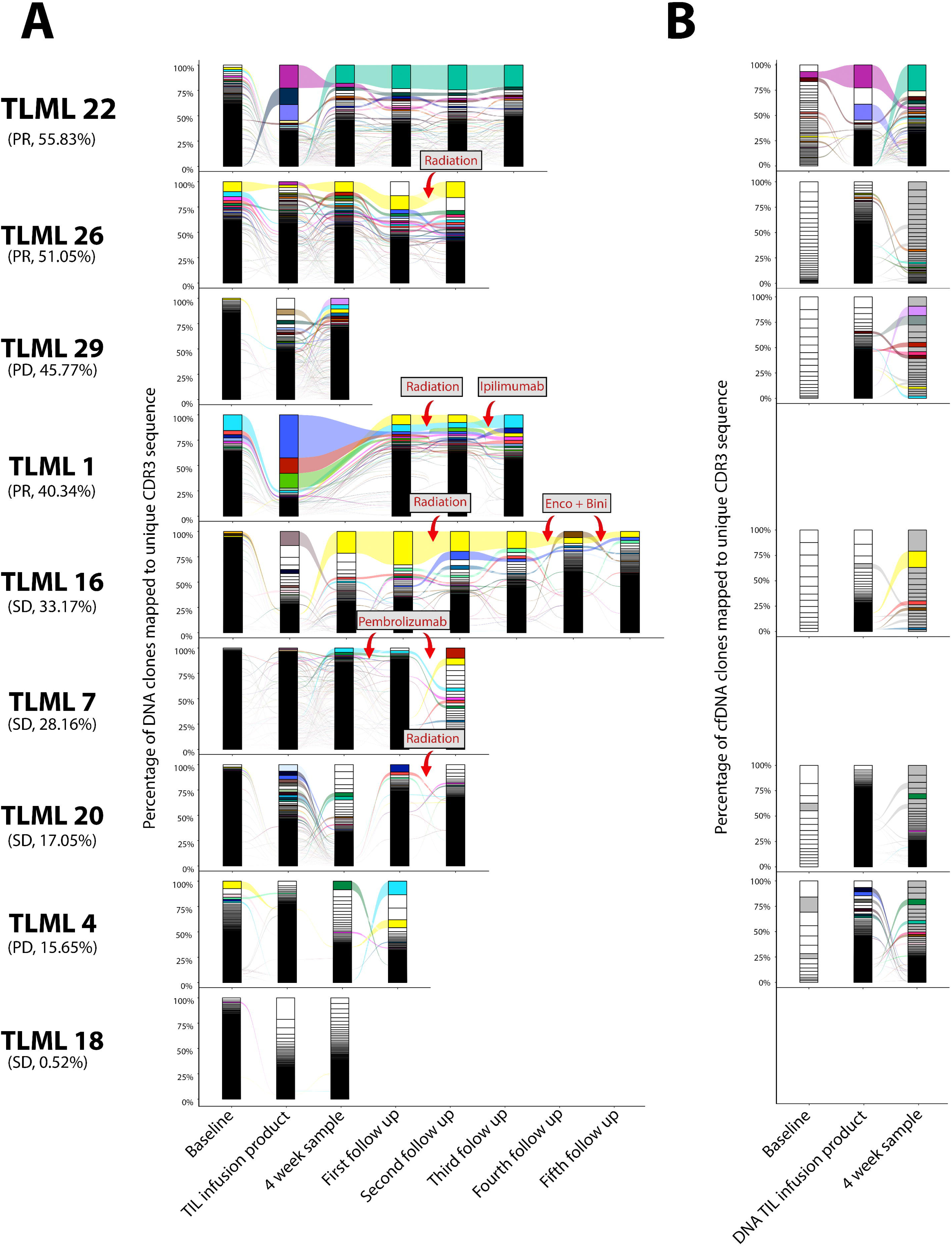
Longitudinal clonotype tracking of tumour-infiltrating lymphocytes (TIL) across DNA and cfDNA T cell repertoires. Series of stacked bars are temporally sorted, with each sub-segment representing an individual clonotype, varying in size depending on its abundance in the repertoire. Patients are sorted according to the abundance of tumour-infiltrating lymphocyte clonotypes in the first post-infusion peripheral blood mononuclear cell DNA repertoire, as shown below each patient ID along with response according to RECIST criteria. (A) Longitudinal tracking of DNA repertoires. The clonotypes which are in the infusion product and one other sample are coloured and tracked throughout the patient timeline. (B) Longitudinal tracking of circulating free DNA repertoires. The DNA TIL infusion product is inserted for reference. Redundant TIL clonotypes from figure 2A and clonotypes present in the 4 week cfDNA repertoire are coloured.

A median TIL abundance of 8.05% (min: 1.36%, max: 38.12%) was found in the PBMC DNA baseline sample and a median TIL abundance of 33.17% (min: 0.52%, max: 55.83%) was found in the first PBMC DNA post-infusion sample. In terms of cfDNA samples, a median TIL abundance of 15.78% (min: 1.67%, max: 51.38%) was found in the baseline sample and a median TIL abundance of 26.10% (min: 14.01%, max: 68.62%) was found in the first post-infusion sample.

Our findings revealed higher TIL abundance of the first PBMC DNA post-infusion sample in responders when compared with non-responders (51.05%, range: 40.34% - 55.83%, n = 3 vs 22.61%, range: 0.52% - 45.77%, n = 6). This difference was determined to be significant by a two-sample two-tailed t-test (p = 0.035). Responders also demonstrated an increased TIL abundance in the PBMC DNA baseline sample when compared with non-responders (37.77%, range: 24.23% - 38.12%, n = 3 vs 4.42%, range: 1.36% - 15.24%, n = 6). This difference was determined to be significant by a two-sample two-tailed t-test (p = 0.00036).

We did not observe a correlation between TIL abundance in PBMC DNA repertoires and TIL abundance in cfDNA repertoires (R^2^ = 0.40). Furthermore, no significant difference was found for the TIL abundance in cfDNA samples between responders and non-responders in either the baseline or first post-infusion sample based on two-sample two-tailed t-tests (p > 0.05).

Overall, our results demonstrate that the abundance of TILs in the PBMC DNA baseline and first PBMC DNA post-infusion T cell repertoires are associated with clinical response.

### 3.4 Responders demonstrate increased overlap of the DNA T cell repertoire between timepoints

To illustrate the impact of sample overlap between timepoints within the same sample-cohort on clinical response, Moritia’s overlap index was calculated between each timepoint in genomic DNA, RNA, and cfDNA (Figure 2A). This quantification of sample overlap allows us to investigate the stability of a patient’s T cell repertoire during treatment and identify potential disruptions. A patient’s mean Morisita’s overlap index was calculated over all samples in the same sample-cohort. Responders demonstrated an increased mean Morisita’s overlap index in DNA samples compared with non-responders (0.46, range: 0.42 - 0.52, n = 3 vs 0.088, range: 0.0048 - 0.35, n = 6). This result was confirmed using a two-sample two-tailed t-test (p = 0.0022). No significant difference was detected between responders and non-responders in terms of their mean Morisita’s overlap index in RNA or cfDNA samples as determined by two-sample two-tailed t-tests (p > 0.05).

To isolate the samples whose overlap differs between responders and non-responders, the Morisita’s overlap index was calculated between each patient’s PBMC DNA baseline, TIL infusion product, and PBMC DNA post-infusion repertoires. Responders demonstrated an increased overlap between the baseline and first post-infusion sample when compared with non-responders (0.52, range: 0.069 - 0.75, n = 3 vs 0.033, range: 0.0079 - 0.12, n = 6). This result was confirmed using a two-sample two-tailed t-test (p = 0.021). Responders also demonstrated an increased overlap between the TIL infusion product and first post-infusion sample when compared with non-responders (0.16, range: 0.14 - 0.45, n = 3 vs 0.037, range: 0.00010 - 0.16, n = 6). This result was confirmed using a two-sample two-tailed t-test (p = 0.047). No significant difference was detected between responders and non-responders in terms of their overlap between the baseline and TIL infusion product DNA repertoire as determined by a two-sample two-tailed t-test (p > 0.05).

Overall, responders demonstrated an increased overlap between timepoints in DNA samples in comparison with responders. Specifically, the PBMC DNA baseline and DNA TIL infusion product had increased overlap with the first PBMC DNA post-infusion sample in comparison with non-responders.

### 3.5 Clonotype’s presence in PBMC DNA baseline and TIL infusion product are predictors of expansion in post-infusion PBMC DNA repertoires

The clonal expansion of T cells results in expanded populations within the T cell repertoire. Traditionally, the identification of expanded clones in T cell repertoire analysis has relied on selecting an arbitrary threshold, such as considering clonotypes representing >1% of the T cell repertoire [16, 29]. However, this approach has inherent limitations as it lacks a systematic and data-driven method for determining the optimal threshold. Patient’s repertoires vary in diversity making a certain threshold too high or too small for certain patients. To address these limitations, we used change point analysis based on the variance in clonotype abundance to determine the threshold for each unique T cell repertoire.

Background clones typically represent a large set of clonotypes that are not subject to specific antigen-driven expansion. These background clones were therefore predicted to have a similar number of T cells per clonotype because they are influenced by similar factors and environmental conditions which do not induce substantial variations in abundance. On the other hand, expanded clones have a greater number of T cells per clonotype and were expected to exhibit larger variations in clone size due to the influence of multiple factors such as antigen density, antigen presentation, and immune stimulation.

Rank abundance curves of clonotypes in a repertoire present a sharp decrease separating these two sub populations. Therefore, we employed the At Most One Change change point algorithm based on variance to the rank abundance curve for clonotypes to identify the point at which the variance in clone size changes the most in a T cell repertoire. Clonotypes above this threshold were labelled as expanded and those below were labelled as background clones. This approach is tailored to PBMC DNA data, as the rationale relies on the representation of individual T cells per clonotype. The median threshold clone percentage for the PBMC baseline, TIL infusion product, and PBMC first post-infusion sample was 0.47% (min: 0.11%, max: 1.61%, n = 27). The corresponding median clone count threshold was 194 (min: 75, max: 1320, n = 27). Expanded clones took up a median of 46.88% of the repertoire (min: 30.90%, max: 82.68%, n = 27). Responders demonstrated a larger amount of the baseline sample taken up by expanded clones (39.41%, range: 36.19% - 40.20%, n = 3 vs 32.38%, range: 30.90% - 37.34%, n = 6). This result was confirmed using a two-sample two-tailed t-test (p = 0.010), however, this higher percentage might not be practically significant. The threshold clone percentage, threshold clone count, and abundance of expanded clones did not differ between responders and non-responders in the TIL infusion product or first post-infusion samples according to two-sample two-tailed t-tests (p > 0.05).

Responders demonstrated a higher proportion of post-infusion expanded clonotypes present in the baseline repertoire (42.86%, range: 38.24% - 48.00%, n = 3 vs 15.21%, range: 6.45% - 33.33%, n = 6). This result was determined to be significant using a two-sample two-tailed t-test (p = 0.0025). Responders also demonstrated a higher percentage of post-infusion expanded clonotypes present in the TIL infusion product (67.86%, range: 60.00% - 76.47%, n = 3 vs 27.60%, range: 0.00% - 61.11%, n = 6). This result was determined to be significant using a two-sample two-tailed t-test (p = 0.019).

To investigate the origin of post-infusion expanded clonotypes, relative risk analysis was conducted on the baseline, TIL infusion product, and first post-infusion DNA repertoires. A clonotype’s presence in the baseline DNA repertoire was found to be a risk factor for that clonotype to expand post-infusion in all patients. All estimated 95% CIs for the calculated risk ratios were found to be greater than 1 for the baseline repertoire compared to the expanding clonotypes in the first post-infusion sample. Additionally, responders were found to have an increased risk ratio (10.61, range: 7.35 - 10.80, n = 3 vs 5.51, range: 3.94 - 5.76, n = 6). This result was confirmed using a two-sample two-tailed t-test (p = 0.0013). Similarly, a clonotype’s presence in the TIL infusion product was found to be a risk factor for that clonotype to expand post-infusion in 7 out of 8 applicable patients. The 7 95% CIs were found to be greater than 1 for the TIL infusion product compared to the expanding clonotypes in the first post-infusion sample. Responders were found to have an increased risk ratio (16.63, range: 3.70 - 20.71, n = 3 vs 2.92, range: 0.78 - 5.99, n = 5). This result was verified by a two-sample two-tailed t-test (p = 0.039). However, it is important to note that the observed range in responders was large.

In summary, the results suggest that clonotypes overlapping in the baseline PBMC repertoire and the TIL infusion product are more likely to expand post-infusion, particularly in responders. As well, responders demonstrate a higher abundance of expanded clonotypes in the baseline and TIL infusion product.

### 3.6 Baseline DNA and cfDNA diversity statistics are associated with clinical response

To demonstrate the effect of diversity of the baseline, TIL infusion product, and post-infusion T cell repertoires on clinical response, multiple diversity statistics were analysed (Figure 4). Clonality and the percentage of the repertoire taken up by large/hyper-expansive clonotypes were chosen as a proxy for clone size skewing in the repertoire. Inverse Simpson diversity, Shannon diversity, and richness were chosen for their representation of overall repertoire diversity.

**Figure 4.**
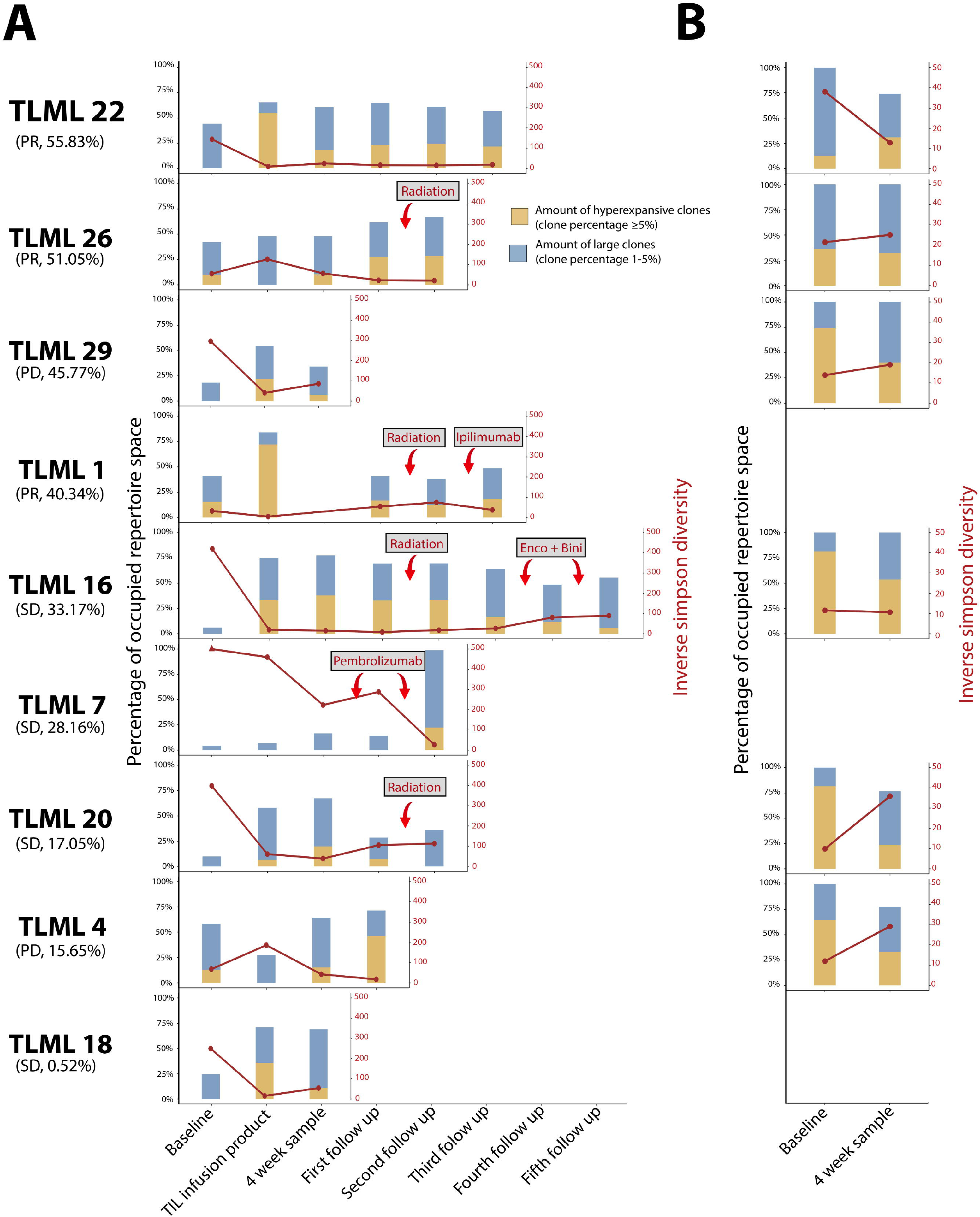
Longitudinal tracking of T cell repertoire diversity in DNA and cfDNA. The line displays the inverse Simpson’s diversity while the stacked bar visualizes the abundance of hyperexpanded and large clonotypes. Hyperexpanded clonotypes were determined to be clonotypes which take up > 5% of the repertoire while large clonotypes were determined to be clonotypes which take up between 1 and 5% of the repertoire. Patients are sorted according to the abundance of tumour-infiltrating lymphocyte clonotypes in the first post-infusion peripheral blood mononuclear cell DNA repertoire, as shown below each patient ID along with response according to RECIST criteria. (A) Longitudinal tracking of the DNA repertoires. (B) Longitudinal tracking of the cfDNA repertoires.

Responders demonstrated an increased clonality of the PBMC baseline DNA repertoire (0.15, range: 0.073 - 0.17, n = 3 vs 0.058, range: 0.048 - 0.095, n = 6). This result was confirmed using a two-sample two-tailed t-test (p = 0.016). Since the percentage of the DNA repertoire taken up by large (between 1% and 5% of T cell repertoire) and hyperexpansive (>5% of T cell repertoire) clones is correlated with the clonality of the repertoire (R^2^ = 0.82), responders also demonstrated an increased percentage of the DNA baseline repertoire taken up by large and hyperexpansive clonotypes (34.60%, range: 25.41% - 35.39%, n = 3 vs 6.17%, range: 1.09% - 31.97%, n = 6). This result was confirmed by a two-sample two-tailed t-test (p = 0.016). No significant difference was found in inverse Simpson diversity, Shannon diversity or richness of the baseline DNA repertoire between responders and non-responders under two-sample two-tailed t-tests (p > 0.05).

Responders demonstrated an increased inverse Simpson diversity, Shannon diversity, and richness of the baseline cfDNA repertoire when compared with non-responders. Under two-sample two-tailed t-tests, the three diversity statistics demonstrated p < 0.05 (p = 0.027, p = 0.0081, p = 0.010 respectively).

Additionally, responders demonstrated a decreased percentage of the baseline cfDNA repertoire taken up by hyperexpansive clones (24.44%, range: 12.71% - 36.18%, n = 2 vs 72.71%, range: 64.18% - 81.54%, n = 4). This result was confirmed by a two-sample two-tailed t-test (p = 0.0057). No association was observed between clonality of the cfDNA baseline repertoire and response under a two-sample two-tailed t-test (p > 0.05).

No significant difference was found in inverse Simpson diversity, Shannon diversity, clonality, percentage of large and hyperexpansive clones or richness of the TIL infusion product or post-infusion DNA repertoires between responders and non-responders under two-sample two-tailed t-tests (p > 0.05). Likewise, no significant difference was found in inverse Simpson diversity, Shannon diversity, clonality, percentage of hyperexpansive clones or richness of the post-infusion cfDNA repertoire between responders and non-responders under two-sample two-tailed t-tests (p > 0.05).

Overall, increased clonality of the baseline DNA repertoire and increased overall diversity of the baseline cfDNA repertoire was associated with response.

## 4. DISCUSSION

In this study, we aimed to evaluate the tracking efficacy of CapTCR-seq for TIL ACT and explored T cell repertoire statistics that could predict and monitor response. We enrolled nine solid tumour patients who received TIL ACT following chemotherapy-induced lymphodepletion. Peripheral blood samples were collected before and after TIL infusion. DNA and RNA were extracted from PBMCs and the TIL infusion product, and cfDNA was extracted from plasma. These samples underwent CapTCR-seq and subsequent T cell repertoire analysis.

One notable observation in our study was the increased overlap between DNA, RNA, and cfDNA repertoires post-infusion. This observation suggests that clonotypes need to reach a specific threshold of T cell abundance to be detected across different sample cohorts. As clonotypes undergo clonal expansion post-infusion, the number of T cells within a given clonotype increases, thereby enhancing the probability of detection across all three sample types. Notebly, we observed a higher overlap in TIL clonotypes between DNA and cfDNA post-infusion. This observation aligns with the notion that TIL clonotypes are more likely to be tumour-reactive and are therefore more likely to expand, thereby increasing T cell turnover. The association between expansion and T cell turnover aligns with a previous study on patients treated with checkpoint inhibitors, which demonstrated a correlation between clonal expansion of a peripheral T cell subset and T cell turnover [27].

Consistent with previous research, we found that the persistence of TILs following infusion was associated with a favourable clinical response. Our study defined persistence as the overall percentage of the post-infusion repertoire composed of TIL clonotypes, which differs from previous studies that focused on the percentage of shared clonotypes between the TIL infusion product and the first PBMC DNA post-infusion sample [14, 20, 22]. Nevertheless, our findings support the notion that a higher proportion of post-infusion samples dominated by TIL clones is indicative of a stronger response. Furthermore, we observed that a higher percentage of the PBMC DNA baseline sample occupied by TIL clonotypes was associated with an enhanced response to TIL ACT. These shared clonotypes between the TIL infusion product and an peripheral sample are likely to represent tumour-reactive clones. Therefore, an increased presence of tumour-reactive clones within a sample is expected to elicit a more potent antitumour response.

Interestingly, responders demonstrated a higher overlap between baseline and post-infusion DNA samples, indicating the persistence of specific T cell clones after treatment despite chemodepletion. Successful treatments seemed to enhance the clonotypes already present in the patient’s immune repertoire, rather than completely replacing the immune system. This finding illuminates the possibility of the persistence of certain T cell subsets which exhibit chemoresistant characteristics, such as less differentiated T cells including naive T cells, stem memory T cells, and memory T cells [9, 10]. These observations support the hypothesis that expanding clones can arise from chemotherapy-resistant baseline clonotypes as well as clonotypes introduced through the TIL infusion product.

To identify expanding clonotypes within PBMC DNA T cell repertoires, we developed a change point analysis method, which provides a data-driven alternative to the arbitrary threshold selection commonly observed in the field [16, 29]. Our results indicated that expanding clones were more abundant in the baseline samples of responders suggesting that the presence of expanding clonotypes in the baseline may serve as an indicator of a favourable response to TIL ACT. Importantly, a clonotype’s presence in the PBMC baseline DNA repertoire and the TIL infusion product were predictors of expansion in post-infusion PBMC DNA repertoires. This finding further supports the hypothesis that baseline clonotypes resistant to chemotherapy can contribute to clonal expansion, alongside clonotypes introduced through TIL infusion. In order to further validate the change point method, future studies could incorporate flow cytometry analysis to identify cell surface markers associated with clonal expansion. By combining flow cytometry with the change point analysis method developed in our study, we can gain valuable insights into the potential of change point analysis to identify expanding clonotypes.

The characteristics of the T cell repertoire, such as diversity and clonality, can impact immune responses. However, their specific association with the response to the ACT of TILs is still not well understood. A previous study on ACT of TILs indicated that the diversity of the TIL infusion product had no effect on the response [14]. Our results support this finding by demonstrating that the richness, diversity, and clonality of the TIL infusion product are not associated with the response to ACT of TILs.

When it comes to analysing the T cell repertoire in the peripheral blood, most studies have focused on melanoma patients treated with checkpoint inhibitors. These studies have revealed that a low diversity of the baseline T cell repertoire in the peripheral blood is correlated with a positive response to anti-DP-1 or anti-CTLA-4 therapy [18]. Our study’s results replicated this observation by showing that an increased clonality of the baseline DNA repertoire in PBMCs and an increased diversity of the cfDNA repertoire are associated with increased response to the ACT of TILs. Increased clonality in the DNA repertoire may indicate clonal expansion, while increased diversity of the cfDNA repertoire may suggest a heightened T cell turnover. These characteristics may be indicative of healthier immune response prior to treatment in these patients.

Regarding post-treatment samples, an increase in richness has been linked to an improved response to anti-CTLA4 therapy [13, 21]. However, in our study, post-infusion PBMC DNA or cfDNA repertoires did not exhibit differences in diversity, richness, or clonality between responders and non-responders.

The small sample size and the inclusion of a specific patient population (melanoma patients) limit the generalizability of our findings. Future studies would benefit from a larger and more diverse cohort alongside the integration of complementary approaches such as flow cytometry and single-cell analysis for a more functional description of the T cell repertoire.

## 5. CONCLUSION

CapTCR-seq was able to track the adoptive cell transfer of TILs with accuracy from PBMC DNA and cfDNA. Increased presence of TILs in the T cell repertoire from DNA extracted from PBMCs both pre- and post-infusion correlated with clinical response according to RECIST criteria. Responders exhibited greater overlap between pre- and post-infusion DNA samples, suggesting the persistence of T cell clonotypes from pre-infusion samples after infusion, despite chemo-depletion.

## 6. LIST OF ABBREVIATIONS

ACT - Adoptive cell transfer; CDR3 - Complementary determining region 3; cfDNA - Circulating free DNA; GAM - Generalized additive model; PBMC - Peripheral blood mononuclear cells; PD - Progressive disease; PR - Partial response; SD - Stable disease; TIL - Tumour-infiltrating lymphocytes

## 7. DECLARATIONS

### 7.1 Ethics approval and consent to participate

The study was approved by the University Health Network Research Ethnics Board under application #11-0683.

### 7.2 Consent for publication

Not Applicable.

### 7.3 Availability of data and materials

The TCR data and scientific computing scripts used to conduct all analysis and to create the figures in this study are in a Github repository at https://github.com/pughlab/CapTCR-TIL-Tracking

### 7.6 Authors’ contributions

CK performed TCR sequence calling, correlative analysis, data interpretation, and drafted the manuscript. SS, DM, AN, and SS supported the CapTCR-seq analysis and interpretation. DG, VS collected and processed blood samples and supported correlative analysis. SP oversaw laboratory work, processed blood samples, performed the CapTCR-seq protocol, and supported data interpretation. LN, NH, and PO supported data interpretation and development of manuscript drafts. MB and TJP conceived and oversaw the study, performed data interpretation, and drafted the manuscript.

## Supporting information

MiXCR Output

Sample Keys

Morisita overlap matrix

Clonotype correlation matrix

## Acknowledgements

We thank the staff of the Princess Margaret Genomics Centre (www.pmgenomics.ca), UHN Bioinformatics and HPC Core (https://bhpc.uhnresearch.ca) for generating and processing the sequencing data used in this study. These programs were enabled through infrastructure support from the Canada Foundation for Innovation, Leaders Opportunity Fund (CFI #32383 and #38401); and the Ontario Ministry of Research and Innovation, Ontario Research Fund Small Infrastructure Program. We also thank past and present members of the Pugh Lab for laboratory and analytical support throughout this project.

