## Supplementary Tables and Figures for "Delineation and monitoring of the T cell repertoire of adoptive cell transfer product during the treatment of advanced melanoma"

**Table S1.** Repertoire taken up by TILs

| | DNA PBMC baseline | DNA PBMC first post-infusion | cfDNA baseline | cfDNA first post-infusion |
|---|---|---|---|---|
| **TLML 1** | 0.37769113 | 0.403444662385469 | | |
| **TLML 4** | 0.152390438247012 | 0.156548938199568 | 0.17910447761194 | 0.140080428954424 |
| **TLML 7** | 0.0297937468114569 | 0.281584682648982 | | |
| **TLML 16** | 0.0804752024085602 | 0.331729173581148 | 0.215384615384615 | 0.330188679245283 |
| **TLML 18** | 0.0136075783744178 | 0.00521785496648385 | | |
| **TLML 20** | 0.0534131623609278 | 0.170532242928006 | 0.0166666666666667 | 0.180257510729614 |
| **TLML 22** | 0.242323290845886 | 0.558338741077223 | 0.513812154696133 | 0.686230248306998 |
| **TLML 26** | 0.381232838553066 | 0.510459679896256 | 0.136518771331058 | 0.191740412979351 |
| **TLML 29** | 0.0349026463319067 | 0.457665472374128 | 0.1125 | 0.390804597701149 |

**Table S2.** DNA PBMC threshold percentage

| | Apheresis / Baseline | TIL Infusion Product | 4 Week sample | FU1 | FU2 | FU3 | FU4 | FU5 |
|---|---|---|---|---|---|---|---|---|
| **TLML 1** | 0.00639938972707239 | 0.00803851745717356 | 0.00489023833194508 | 0.00489023833194508 | 0.00307692307692308 | 0.00670538792085753 | | |
| **TLML 4** | 0.0100224103585657 | 0.00147337699416994 | 0.00533004179874884 | 0.0072681 | | | | |
| **TLML 7** | 0.00110779097733401 | 0.00172510489063687 | 0.00181840060969903 | 0.00244203196402234 | 0.04649682 | | | |
| **TLML 16** | 0.00170877578420603 | 0.00503856807146098 | 0.0160969133753734 | 0.011400037686075 | 0.0130818568616244 | 0.0106837606837607 | 0.00708684286959047 | 0.0080563947633434 |
| **TLML 18** | 0.00392526299262051 | 0.0096254812740637 | 0.0108165752589884 | | | | | |
| **TLML 20** | 0.00198001131435037 | 0.00264154326820285 | 0.00471466237654735 | 0.00328618374053436 | 0.00400393829996718 | | | |
| **TLML 22** | 0.00608342989571263 | 0.00422349110471427 | 0.00618645900930132 | 0.00755187535902932 | 0.00602565763897888 | 0.00508334917918489 | | |
| **TLML 26** | 0.00561104699749294 | 0.00327038831786871 | 0.00491417261031294 | 0.00685308905572009 | 0.0117931312201576 | | | |

| | | | |
|---|---|---|---|
| **TLML 29** | 0.0031346072 6987763 | 0.002274605485 81323 | 0.00393907 012801978 |

**Table S3.** DNA PBMC threshold clone count

| | **Apheresis / Baseline** | **TIL Infusion Product** | **4 Week sample** | **FU1** | **FU2** | **FU3** | **FU4** | **FU5** |
|---|---|---|---|---|---|---|---|---|
| **TLML 1** | 12 | 11 | 28 | 28 | 35 | 18 | | |
| **TLML 4** | 15 | 112 | 31 | 22 | | | | |
| **TLML 7** | 185 | 119 | 82 | 76 | 5 | | | |
| **TLML 16** | 112 | 29 | 9 | 17 | 10 | 13 | 27 | 24 |
| **TLML 18** | 48 | 16 | 19 | | | | | |
| **TLML 20** | 111 | 53 | 32 | 43 | 43 | | | |
| **TLML 22** | 35 | 15 | 25 | 21 | 27 | 31 | | |
| **TLML 26** | 21 | 68 | 34 | 21 | 14 | | | |
| **TLML 29** | 55 | 41 | 36 | | | | | |

**Table S4.** Repertoire taken up by expanded clones

| | **Apheresis / Baseline** | **TIL Infusion Product** | **4 Week sample** | **FU1** | **FU2** | **FU3** | **FU4** | **FU5** |
|---|---|---|---|---|---|---|---|---|
| **TLML 1** | 0.3619257501 2714 | 0.826766261404 581 | 0.41749738 0229465 | 0.41749738 0229465 | 0.4272978303 74753 | 0.4427917079 85078 | | |
| **TLML 4** | 0.3197211155 37849 | 0.468829969475 013 | 0.64295733 1612758 | 0.67819468 4190868 | | | | |
| **TLML 7** | 0.3374681145 68909 | 0.375852791049 332 | 0.34608442 1922424 | 0.34720375 2508523 | 0.3184713375 79618 | | | |
| **TLML 16** | 0.3279628951 544 | 0.748362803990 221 | 0.52240292 0677066 | 0.62788141 4484015 | 0.5032069600 55884 | 0.4481605351 17057 | 0.39541097879 756 | 0.38039610 6075864 |
| **TLML 18** | 0.3193593970 79604 | 0.590676408820 441 | 0.48217550 274223 | | | | | |
| **TLML 20** | 0.3734442768 24439 | 0.644495175562 621 | 0.69276489 2523573 | 0.37493927 7039577 | 0.4295372497 53856 | | | |
| **TLML 22** | 0.3941193511 | 0.673174932754 | 0.55649289 | 0.57535762 | 0.5932875469 | 0.5673195157 | | |

| | | | | | | |
|---|---|---|---|---|---|---|
| | 00811 | 471 | 7829692 | 8234767 | 74213 | 50776 |
| **TLML 26** | 0.402005650841657 | 0.576772637388905 | 0.491587892024707 | 0.56622356911633 | 0.535580739803256 | |
| **TLML 29** | 0.309030080173609 | 0.586572744087206 | 0.407128040731661 | | | |

**Table S5.** Percentage of DNA PBMC post-infusion expanded clones present in repertoire

| | DNA PBMC baseline | DNA TIL infusion product |
|---|---|---|
| **TLML 4** | 0.428571428571429 | 0.678571428571429 |
| **TLML 16** | 0.0645161290322581 | 0.0967741935483871 |
| **TLML 20** | 0.146341463414634 | 0.378048780487805 |
| **TLML 22** | 0.333333333333333 | 0.333333333333333 |
| **TLML 26** | 0.157894736842105 | 0 |
| **TLML 29** | 0.09375 | 0.21875 |
| | 0.48 | 0.6 |
| | 0.382352941176471 | 0.764705882352941 |
| | 0.166666666666667 | 0.611111111111111 |

**Table S6.** Relative risk of reference sample clones expanding in DNA PBMC post-infusion

| | DNA PBMC baseline | DNA TIL infusion product |
|---|---|---|
| **TLML 1** | 10.7954545454545 | 20.7068557919622 |
| **TLML 4** | 5.56650246305419 | 0.781055900621118 |
| **TLML 7** | 5.6390977443609 | 4.68203497615262 |
| **TLML 16** | 4.52941176470588 | 2.92 |
| **TLML 18** | 5.75892857142857 | 0 |
| **TLML 20** | 3.94252873563218 | 2.01209302325581 |
| **TLML 22** | 7.34978229317852 | 3.7007299270073 |

| | | |
|---|---|---|
| **TLML 26** | 10.6091954022989 | 16.6279069767442 |
| **TLML 29** | 5.44545454545455 | 5.99335548172758 |

**Table S7.** Longitudinal data for DNA inverse simpson diversity

| | Apheresis / Baseline | TIL Infusion Product | 4 Week sample | FU1 | FU2 | FU3 | FU4 | FU5 |
|---|---|---|---|---|---|---|---|---|
| **TLML 1** | 32.3469717416468 | 4.41004475264349 | 54.3547204434883 | 54.354720443488<br>3 | 74.1694071307758 | 37.6921199897227 | | |
| **TLML 4** | 67.4305059872995 | 185.546661217418 | 42.1526069280112 | 16.921159953110<br>6 | | | | |
| **TLML 7** | 805.235265455923 | 459.863975752931 | 222.848517814854 | 287.45984348881<br>4 | 26.5248364325069 | | | |
| **TLML 16** | 420.104469241304 | 20.3469987921656 | 15.0984679365188 | 8.634876476771<br>53 | 18.0628232469681 | 26.5930036879229 | 80.9014652589625 | 89.7002921937333 |
| **TLML 18** | 249.719688580108 | 15.8743191212946 | 54.673294294663 | | | | | |
| **TLML 20** | 398.548008118901 | 61.1231056750012 | 38.7992980851603 | 105.50378194846 | 113.190582337237 | | | |
| **TLML 22** | 144.106855966357 | 9.51031507468729 | 24.4790452121271 | 16.086957034838<br>2 | 15.1118643990723 | 18.8683386071158 | | |
| **TLML 26** | 55.1913449980409 | 126.297689923858 | 56.1477579203565 | 22.491430638870<br>9 | 20.5785857559774 | | | |
| **TLML 29** | 295.766835349303 | 41.111735159527 | 85.9482791584076 | | | | | |

**Table S8.** Longitudinal data for DNA shannon diversity

| | Apheresis / | TIL Infusion | 4 Week | FU1 | FU2 | FU3 | FU4 | FU5 |
|---|---|---|---|---|---|---|---|---|

| | Baseline | Product | sample | | | | | |
|---|---|---|---|---|---|---|---|---|
| TLML 1 | 5.02262460447881 | 2.62005052031945 | 5.32118790060495 | 5.32118790060495 | 5.69139355533657 | 4.93583537679784 | | |
| TLML 4 | 4.89475683071017 | 6.46710486114634 | 4.76477224180496 | 4.03196150193797 | | | | |
| TLML 7 | 7.14058046365357 | 6.65049564324609 | 6.52532517712803 | 6.31423128173427 | 3.5130059404662 | | | |
| TLML 16 | 6.70044729644394 | 4.12144806757085 | 3.85517014772385 | 3.81634735877481 | 4.14001069801264 | 4.5040066244045 | 5.19068667916479 | 5.08035691637213 |
| TLML 18 | 5.89577933017533 | 4.06753417974074 | 4.63284372559351 | | | | | |
| TLML 20 | 6.52319402553804 | 5.27867176342891 | 4.64634180335137 | 5.86555511841658 | 5.6893426496096 | | | |
| TLML 22 | 5.37828718170048 | 3.83589317717793 | 4.68779402762784 | 4.3899812813966 | 4.50280222614234 | 4.74684989146852 | | |
| TLML 26 | 5.24037096109265 | 5.62270637011366 | 5.24369221975907 | 4.52977512975388 | 4.20620292009006 | | | |
| TLML 29 | 6.10671690499382 | 5.32487235546 | 5.5858134971439 | | | | | |

**Table S9.** Longitudinal data for DNA richness

| | Apheresis / Baseline | TIL Infusion Product | 4 Week sample | FU1 | FU2 | FU3 | FU4 | FU5 |
|---|---|---|---|---|---|---|---|---|
| TLML 1 | 415 | 450 | 508 | 508 | 845 | 400 | | |
| TLML 4 | 223 | 1738 | 572 | 439 | | | | |
| TLML 7 | 1814 | 1211 | 1288 | 873 | 45 | | | |
| TLML 16 | 1218 | 657 | 171 | 265 | 223 | 238 | 334 | 271 |
| TLML 18 | 518 | 330 | 222 | | | | | |
| TLML 20 | 1025 | 1120 | 704 | 734 | 655 | | | |
| TLML 22 | 330 | 815 | 475 | 419 | 509 | 587 | | |
| TLML 26 | 478 | 817 | 526 | 428 | 230 | | | |
| TLML 29 | 657 | 1367 | 621 | | | | | |

**Table S10.** Longitudinal data for DNA clonality

| | Apheresis / Baseline | TIL Infusion Product | 4 Week sample | FU1 | FU2 | FU3 | FU4 | FU5 |
|---|---|---|---|---|---|---|---|---|
| **TLML 1** | 0.16682273593978 | 0.571133681386357 | 0.145942742085676 | 0.145942742085676 | 0.155496472422323 | 0.176188836971369 | | |
| **TLML 4** | 0.0947657966877532 | 0.13315283633615 | 0.249540410396737 | 0.337338829670431 | | | | |
| **TLML 7** | 0.04834001949474 | 0.0632051484822902 | 0.0887493932727724 | 0.0675883977824689 | 0.0771427858638034 | | | |
| **TLML 16** | 0.0569345698823073 | 0.364727373307935 | 0.250209566347798 | 0.316033665106668 | 0.234348218811126 | 0.176940087032892 | 0.10676979177786 | 0.0931365294436294 |
| **TLML 18** | 0.0566715704230696 | 0.298591275893465 | 0.142491139460189 | | | | | |
| **TLML 20** | 0.0590345390885634 | 0.24816854629891 | 0.291368176417373 | 0.111078715967421 | 0.122642609356846 | | | |
| **TLML 22** | 0.0725640195516404 | 0.427750928009784 | 0.239403766207252 | 0.272925615731246 | 0.277522698274423 | 0.255398994415756 | | |
| **TLML 26** | 0.150615624176269 | 0.161495826039375 | 0.163058240663936 | 0.25240418728292 | 0.226527845375808 | | | |
| **TLML 29** | 0.058721588845163 | 0.262521238391166 | 0.1314685209046 | | | | | |

**Table S11.** Longitudinal data for abundance of large clones (>1% of DNA repertoire)

| | Apheresis / Baseline | TIL Infusion Product | 4 Week sample | FU1 | FU2 | FU3 | FU4 | FU5 |
|---|---|---|---|---|---|---|---|---|
| **TLML 1** | 0.346033225970504 | 0.784519269263996 | 0.298438885455571 | 0.298438885455571 | 0.30172255095332 | 0.39443736128819 | | |
| **TLML 4** | 0.319721115537849 | 0.162233611324559 | 0.56269812326423 | 0.576299930347355 | | | | |
| **TLML 7** | 0.0109029954084979 | 0.0315275342358779 | 0.09208327205145 | 0.0623564399526101 | 0.931210191082803 | | | |
| **TLML 16** | 0.0557996663818707 | 0.659544497192895 | 0.585794888815134 | 0.638464920545192 | 0.534832031498063 | 0.448160535117057 | 0.273308161487075 | 0.309097012420275 |

| | | | | | | |
|---|---|---|---|---|---|---|
| **TLML 18** | 0.0718584811849061 | 0.581050927546377 | 0.48217550274223 | | | |
| **TLML 20** | 0.0346973411276636 | 0.435930506031409 | 0.620868153312181 | 0.193199028432633 | 0.273952521605951 | |
| **TLML 22** | 0.254128041714948 | 0.632444198008589 | 0.485081837190857 | 0.533871351911787 | 0.511377478294674 | 0.460797363250301 |
| **TLML 26** | 0.353854112778065 | 0.289054520958734 | 0.364706685322322 | 0.511433294304015 | 0.558131507794972 | |
| **TLML 29** | 0.0676653203930315 | 0.464538979182244 | 0.270517736291826 | | | |

**Table S12.** Longitudinal data for abundance of hyperexpanded clones (>5% of DNA repertoire)

| | **Apheresis / Baseline** | **TIL Infusion Product** | **4 Week sample** | **FU1** | **FU2** | **FU3** | **FU4** | **FU5** |
|---|---|---|---|---|---|---|---|---|
| **TLML 1** | 0.155661976606204 | 0.722729092479825 | 0.166160625520595 | 0.166160625520595 | 0.128336620644313 | 0.179864947820749 | | |
| **TLML 4** | 0.128112549800797 | 0 | 0.152733750385727 | 0.458970554091841 | | | | |
| **TLML 7** | 0 | 0 | 0 | 0 | 0.222292993630573 | | | |
| **TLML 16** | 0 | 0.330852425488111 | 0.379770992366412 | 0.32943910558382 | 0.335365466438052 | 0.166945373467113 | 0.118268951495789 | 0.057267539442766 |
| **TLML 18** | 0 | 0.359730486524326 | 0.108622790981109 | | | | | |
| **TLML 20** | 0 | 0.065114386410191 | 0.197487003023938 | 0.0721817402486069 | 0 | | | |
| **TLML 22** | 0 | 0.547667641923458 | 0.179191001514168 | 0.23169826124725 | 0.243890112738111 | 0.216416302212081 | | |
| **TLML 26** | 0.098810139679255 | 0 | 0.103641265399447 | 0.274950065431504 | 0.284185698671115 | | | |
| **TLML 29** | 0 | 0.218708433355633 | 0.0636956020701071 | | | | | |

**Table S13.** Longitudinal data for cfDNA inverse simpson diversity

| | Apheresis / Baseline | 4 Week sample |
|---|---|---|
| TLML 4 | 11.9706667 | 29.1350566 |
| TLML 16 | 11.5753425 | 10.7009524 |
| TLML 20 | 9.89010989 | 35.8749575 |
| TLML 22 | 38.0499419 | 12.8411865 |
| TLML 26 | 21.4247567 | 25.0427108 |
| TLML 29 | 13.8079827 | 18.9699248 |

**Table S14.** Longitudinal data for cfDNA shannon diversity

| | Apheresis / Baseline | 4 Week sample |
|---|---|---|
| TLML 4 | 2.66458144 | 4.01860794 |
| TLML 16 | 2.53288508 | 2.7031899 |
| TLML 20 | 2.45709667 | 4.12386192 |
| TLML 22 | 3.79861526 | 3.84631048 |
| TLML 26 | 3.25677165 | 3.41350726 |
| TLML 29 | 2.73379632 | 3.08241232 |

**Table S15.** Longitudinal data for cfDNA richness

| | Apheresis / Baseline | 4 Week sample |
|---|---|---|
| TLML 4 | 17 | 156 |
| TLML 16 | 14 | 20 |
| TLML 20 | 15 | 163 |
| TLML 22 | 51 | 143 |
| TLML 26 | 33 | 39 |
| TLML 29 | 18 | 25 |

**Table S16.** Longitudinal data for cfDNA clonality

| | Apheresis / Baseline | 4 Week sample |
|---|---|---|
| TLML 4 | 0.05951966 | 0.20421336 |
| TLML 16 | 0.04023113 | 0.09765304 |
| TLML 20 | 0.09266945 | 0.19040751 |
| TLML 22 | 0.03388003 | 0.22497866 |
| TLML 26 | 0.06856439 | 0.06825445 |
| TLML 29 | 0.05417138 | 0.04239477 |

**Table S17.** Longitudinal data for abundance of hyperexpanded clones (>5% of cfDNA repertoire)

| | Apheresis / Baseline | 4 Week sample |
|---|---|---|
| TLML 4 | 0.64179104 | 0.33009383 |
| TLML 16 | 0.81538462 | 0.53773585 |
| TLML 20 | 0.71666667 | 0.23440079 |
| TLML 22 | 0.12707182 | 0.31226486 |
| TLML 26 | 0.36177474 | 0.32448378 |
| TLML 29 | 0.7375 | 0.40229885 |

**Figure S18.** Change point plots

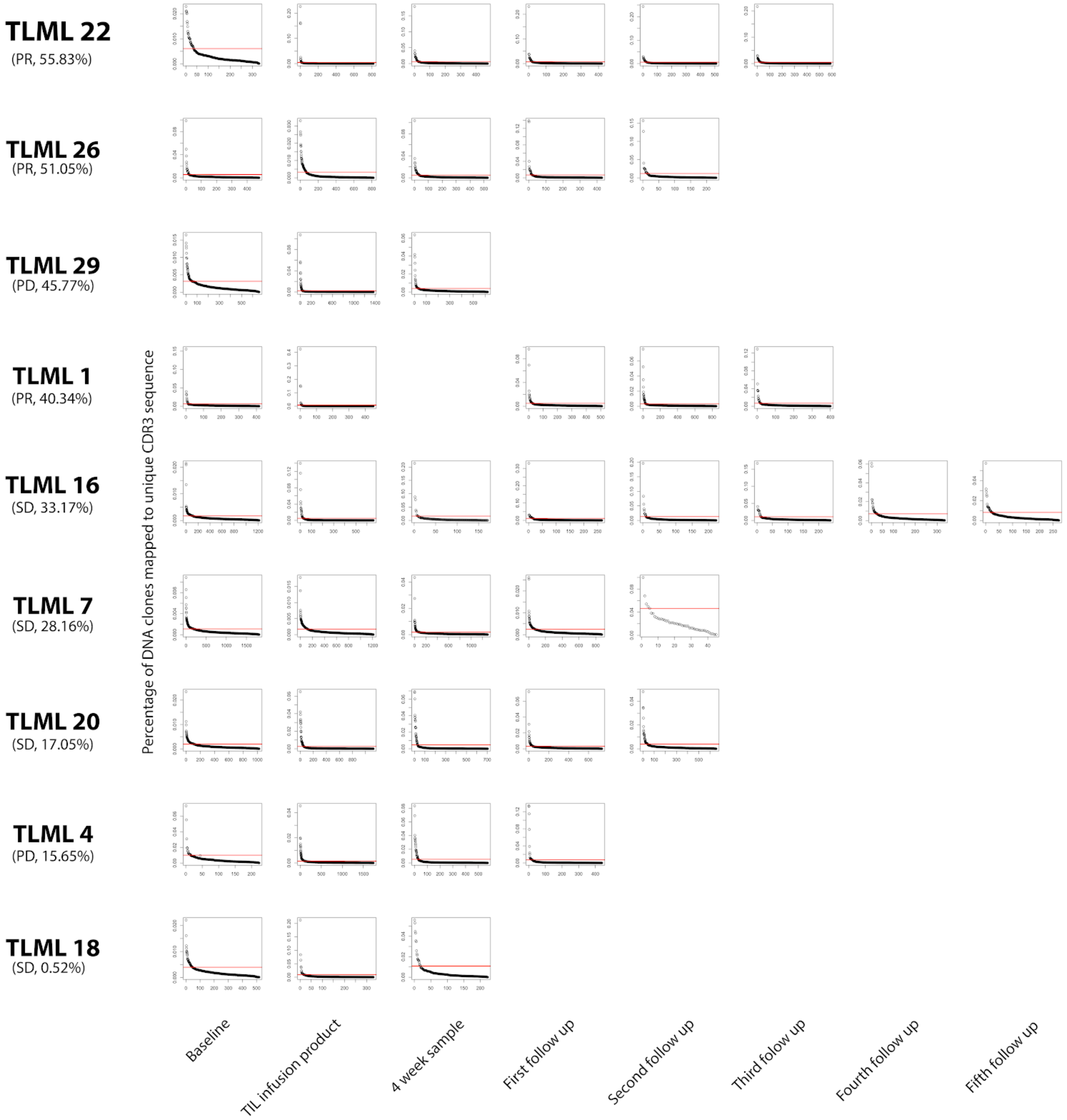

TLML 22
(PR, 55.83%)
TLML 26
(PR, 51.05%)
TLML 29
(PD, 45.77%)
TLML 1
(PR, 40.34%)
TLML 16
(SD, 33.17%)
TLML 7
(SD, 28.16%)
TLML 20
(SD, 17.05%)
TLML 4
(PD, 15.65%)
TLML 18
(SD, 0.52%)
Percentage of DNA clones mapped to unique CDR3 sequence
Baseline
TIL infusion product
4 week sample
First follow up
Second follow up
Third follow up
Fourth follow up
Fifth follow up

**Table S19.** Clone tracking circle plot

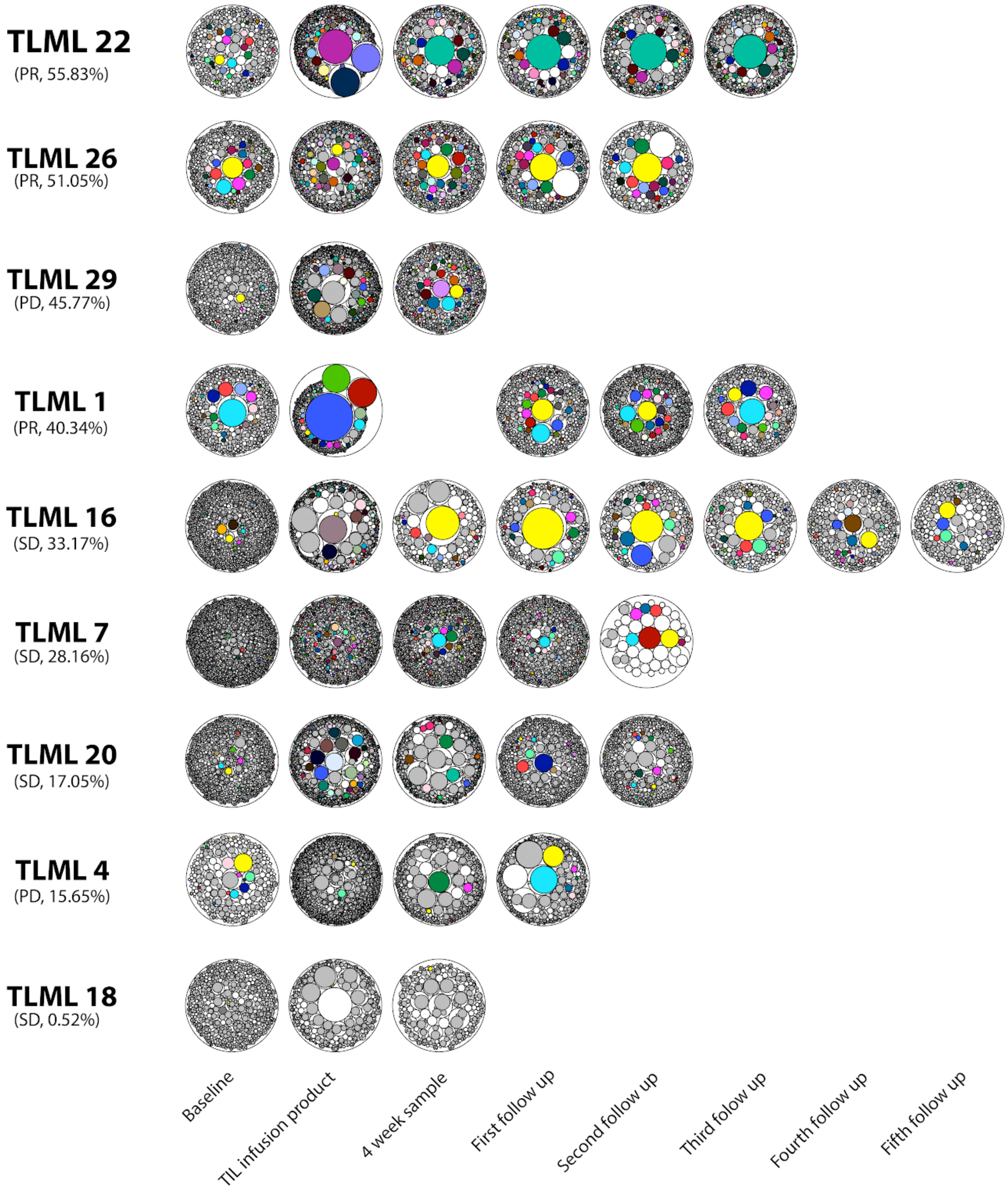
